# Pharmacoepidemiology of Oxycodone in the USA: an observational study of ARCOS, Medicaid, and Medicare drug databases

**DOI:** 10.1101/2025.05.15.25327730

**Authors:** Jay P. Solgama, Apoorva Pradhan, Kenneth L. McCall, Brian J. Piper

## Abstract

The US continues to battle an opioid crisis, substantially influenced by prescription opioids, with nearly 80,000 opioid-related deaths in 2023. This study aims to characterize oxycodone’s distribution in the United States (US) from 2000-23 using three data sources. Morphine Milligram Equivalents (MME) of oxycodone were calculated from the Drug Enforcement Administration annual summary reports from the Automation of Reports and Consolidated Orders System (ARCOS); the number of units and prescriptions of oxycodone per enrollee were calculated using the Medicaid State Drug Utilization Data (M-SDUD); and the number of claims and 30-day fills per enrollee and per beneficiary with a claim for oxycodone were calculated from the Medicare Part D Prescribers (M-PDP) dataset. Oxycodone MME per person in ARCOS rose +280% from 2000 to 2010 before declining, ending at +56% over the full period. At the 2010 peak Florida, Delaware, and Tennessee showed high distributions of oxycodone, while Texas and Illinois had lower amounts compared to most states. Delaware again stood out in the Medicaid data with higher numbers of units and prescriptions, along with Alaska, Arizona, Maine, and Maryland. This data also had some anomalies in the number of prescriptions in the 2000s. The Medicare data covered 2013-22, with the District of Columbia and North Dakota standing out with lower numbers of claims and 30-day fills per beneficiary compared to other states. Oxycodone distribution varied substantially between states over time. Each dataset provided complimentary insights, highlighting the importance of multi-source monitoring for assessing opioid distribution trends and informing public policy.

## INTRODUCTION

The United States (US) has been battling an opioid crisis for decades and in 2023 a staggering 96.4% of opioid misusers were using prescription pain relievers (not including illegally made fentanyl).^1^ Prescription opioids account for the vast majority of opioid misuse, with oxycodone standing out as a major contributor due to marketing practices and high abuse liability.^2–4^ The US healthcare system plays a central role in both perpetuating and combating the epidemic, which claimed 79,358 lives in 2023 as reported by the National Center for Health Statistics.^5^ This study aims to characterize the distribution of the widely used prescription opioid oxycodone throughout the US by leveraging three data sources that provide state-level data: the US Drug Enforcement Administration Automated Reports and Consolidated Ordering System (ARCOS), the Medicaid State Drug Utilization Data (M-SDUD), and the Medicare Part D Prescribers dataset (M-PDP). ARCOS is a comprehensive database that covers the entire US population. Medicaid is responsible for 12-28% of the US population while Medicare Part D covers 12-16% of the US population. Each data source provides different, but possibly complementary, information that can be utilized to monitor prescription drug distribution.

Studies examining global consumption of opioids using International Narcotics Control Board (INCB) data and the IQVIA Midas® database have revealed the US as the third highest consumer of opioids in 2015-17 and 2019.^6,7^ These reports also show that oxycodone is the most heavily consumed opioid. Within the US, 31.3% of people who misused prescription opioids were misusing oxycodone products (including generic oxycodone and brand name formulations such as OxyContin, Percocet, Percodan, and Roxicodone), second to only hydrocodone at 42.8%.^1^ Oxycodone’s abuse liability is distinguished from other opioids by its markedly higher desirability and milder side-effect profile, supported by studies of its unique pharmacologic properties.^2,3^

ARCOS is an infrequently utilized database in the literature, particularly at the level of individual drugs. Previous studies have used ARCOS to characterize oxycodone distribution from 2000-21, showing a substantial increase in distribution to a peak in 2010, followed by a slower decline until 2021.^8,9^ Another report looked at the subset of ARCOS data released by the Washington Post in the context of commuting zones which showed a superlinear scaling relationship between population size and amount of oxycodone, as well as regions of higher distribution in the Appalachians, Ozarks, and the west coast.^10^ Other investigations have examined ARCOS oxycodone distribution in specific states and territories, including Florida, Delaware, Maryland, Virginia, and Puerto Rico.^11–13^

The Medicaid and Medicare programs started in 1965 to provide health insurance to Americans.^14^ Medicaid is a government program, administered by states according to federal guidelines, that provides health coverage for people with low income or limited resources.^15^ Medicaid services can vary by state, but must fall within the parameters set by the federal government. Medicare is a federal program that provides health coverage for individuals over age 65, with certain disabilities, or with end-stage renal disease.^16^ Both programs together offer health insurance for millions of Americans who otherwise may not have the resources to get coverage.

M-SDUD and M-PDP datasets have been underutilized for studying oxycodone-specific trends, though they may offer valuable insights into opioid prescription patterns. Some reports have examined opioids broadly and medications for opioid use disorder, including buprenorphine and naltrexone, using the M-SDUD.^17–25^ The M-PDP has been used to evaluate opioid prescribing trends by medical oncologists.^26^ Both databases have been just as infrequently used to examine drugs in other classes. There is a wealth of information included in these databases that can be used to characterize trends in opioids as well as a broad range of other drugs. However, prior reports employing the M-SDUD have also noted some anomalous data points which needed to be addressed prior to analysis.^17,27,28^

This report investigates oxycodone distribution using the ARCOS, M-SDUD, and M-PDP data with the most recent information available. ARCOS overcomes limitations in the frequently used IQVIA database, which has gaps in Veteran’s Affairs, Indian Health Services facilities, hospitals, and independent pharmacies.^29^ All three sources provide data at the state-level, allowing for a more complete picture of oxycodone distribution trends and identifying geographic disparities and potential regulatory impacts.

## METHODS

### Procedures

Data was collected from the ARCOS Report 5, the M-SDUD database, and the M-PDP dataset. Informed consent was not required as all data used is publicly available and deidentified. The Python package pandas (version 2.2.3, RRID:SCR_018214) was used to organize and consolidate the raw data for analysis.^30^

ARCOS is reported by the US Drug Enforcement Administration and includes distribution of Schedule II-IV controlled substances.^31^ The weights for oxycodone were reported annually from 2000-23 for each state. Amounts were provided in grams grouped by business type (hospitals, pharmacies, practitioners, mid-level practitioners, narcotic treatment programs, teaching institutions). Pharmacies, hospitals, practitioners, and the sum of all business types were examined in this report. Data was read from the PDF reports using the Python package tabula-py (version 2.10.0) and filtered for oxycodone.^32^

M-SDUD is provided by the Centers for Medicare and Medicaid Services (CMS) and includes all drugs provided by manufacturers who participate in the Medicaid Drug Rebate program.^33^ Drug utilization data was reported quarterly per state and grouped by the NDC code for each drug from 2000-23. Units reimbursed (based on the unit type of the NDC code), number of prescriptions, and reimbursement amounts are all provided. Counts under 11 are suppressed to protect individual privacy. Data was filtered for oxycodone using the NDC codes, brand names, and generic names provided in the NDC directory.^34^ Data was also searched for text stems specific for oxycodone or its brand names (e.g., “oxyco”, “percoc”, “endoce”, etc.) to account for misspellings or truncations present in the data.

M-PDP data is provided by the CMS and includes a dataset reporting the geography and the drug.^35^ The data was reported per state from 2013 (i.e. the earliest year available) to 2022 by the brand and generic names for oxycodone formulations. Data included number of prescribers, total claims, total 30-day fills, total drug cost, and number of beneficiaries. The same fields were also provided for beneficiaries over 65 years old, as well as the cost share for beneficiaries with and without low-income subsidies. Claims fewer than 11 were suppressed to protect individual privacy. Data was filtered for oxycodone similarly to the Medicaid data as above.

### Data analysis

Microsoft Excel was used to analyze the data after aggregation and formatting. RStudio (version 2024.12.0+467, RRID:SCR_000432) was used to visualize the data using the tidyverse (version 2.0.0, RRID:SCR_019186) collection of packages.^36^

A standard oral conversion factor of 1.5 was used to convert grams of oxycodone to morphine milligram equivalents (MME) for the weights provided in ARCOS. ^9^ MMEs were then used to calculate MME per buyer and MME per person by dividing the values by the number of businesses that made purchases and the state population reported by the U.S. Census Bureau American Community Survey, respectively.

Units and prescriptions per enrollee were calculated for the Medicaid data by dividing each value by the reported number of enrollees for each state. Similarly, the number of Medicare Part D enrollees was used to calculate claims, 30-day fills, and cost per enrollee. Each field was also divided by the number of beneficiaries with claims for each particular drug to get claims, 30-day fills, and cost per beneficiary.

Values were plotted across the time for each state to characterize the distribution of oxycodone per population unit. The five states with the highest average distribution and the five states with the lowest average distribution were plotted for pharmacies, hospitals, and the total of all business types in ARCOS; units and prescriptions per enrollee for Medicaid; and claims and 30-day fills both per enrollee and per beneficiary for Medicare. Data for ARCOS practitioners varied widely, and the ten states with the highest distribution were plotted. Extreme values in the Medicaid data (greater than 5*IQR+Q3) are shown separately.

For each set of values, the range of ± standard deviations was identified and included as a shaded region on each plot. A scatter plot was created, and a Pearson correlation coefficient was calculated, for each unique combination of ARCOS MME/Person, Medicaid Units/Enrollee, Medicare Claims/Enrollee, and Medicare Claims/Beneficiary. Choropleth maps of the U.S. states were created for 2013, the earliest common year between the datasets.

## RESULTS

### Total (i.e. National) Oxycodone Distribution

There were 1,491.16 metric tons (in MME) of oxycodone distributed throughout the US from 2000-23, for an average of 71.0 metric tons per year. The total MME per person increased by +280% from 2000 to a peak in 2010, then gradually fell until 2023 for an overall increase of +56% over the whole time-period. Stratifying the data into business types reveals pharmacies as the largest contributing group (more than 94% of total weight from 2000-23). MME per person of oxycodone distributed by pharmacies increased by +293% from 2000 to a peak in 2010, before falling to +63% of the original value by 2023. Practitioners account for only 0.4% of the total weight of oxycodone distributed in the US and follow a similar trend to pharmacies. There was a sharp increase of +15,565% to a peak in 2010, due to a small number of states, before a decrease to 63.4% of the original value by 2023. Hospitals account for 5.3% of the total weight of oxycodone distributed in the US, and MME per person was substantially less variable between years. MME per person of oxycodone distributed by hospitals increased by +89% from 2000 to a later peak in 2012 and decreased to −13% of the original value by 2023. (Figure 1, Figure 2)

**Figure 1.**
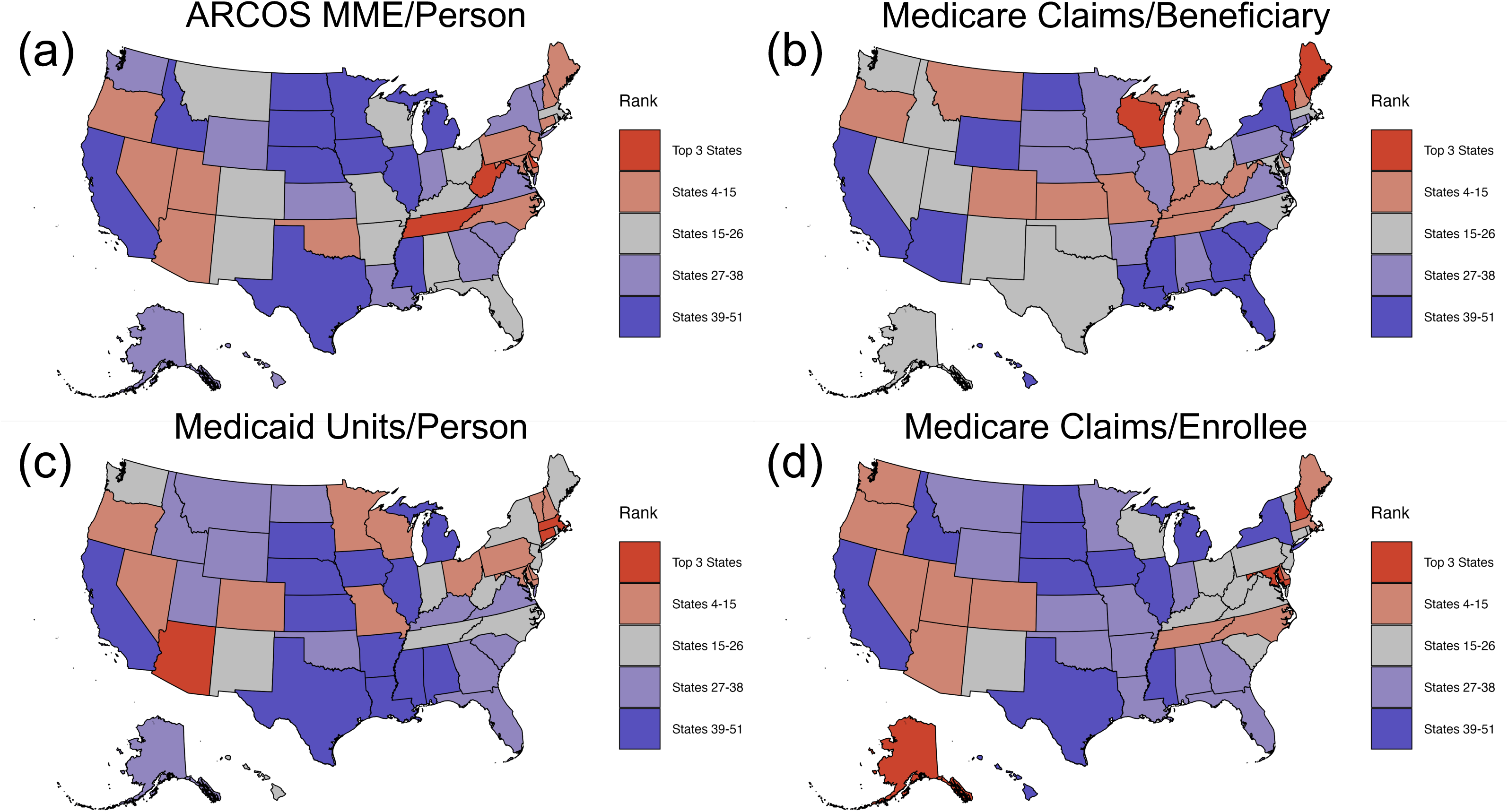
Choropleth maps of the US states ranked by oxycodone distribution in 2013 for **(a)** ARCOS MME per person, **(b)** Medicare Part D Prescribers claims per beneficiary, **(c)** Medicaid SDUD units per enrollee, and (d) Medicare Part D Prescribers claims per enrollee.

**Figure 2.**
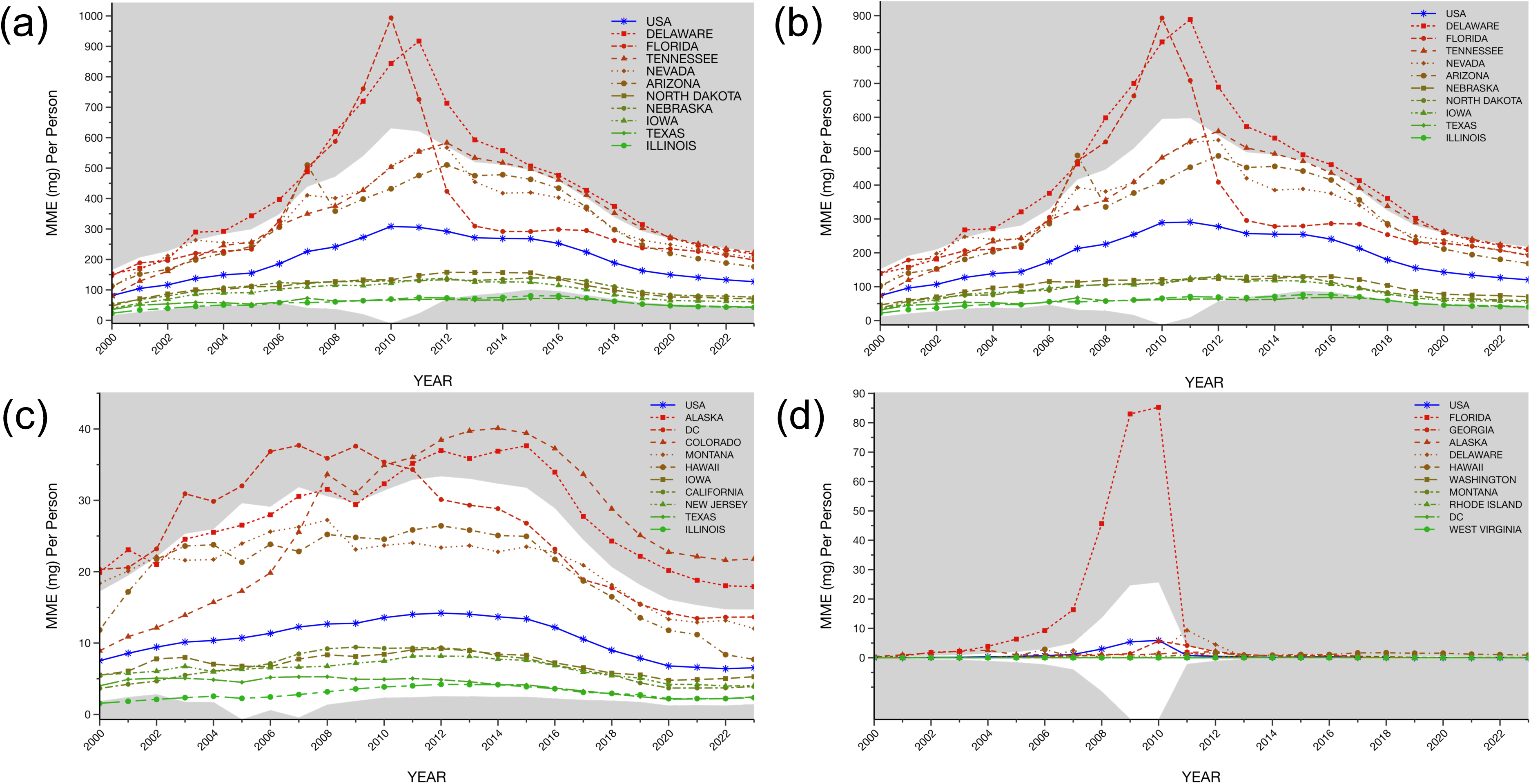
Oxycodone distribution for the top 5 and bottom 5 states (a,b,c) and top 10 states (c) as reported by the Drug Enforcement Administration’s Automated Reports and Consolidated Ordering System in **(a)** total, **(b)** by pharmacies, **(c)** by hospitals, **(d)** and by practitioners. The shaded regions indicate ±1.96 standard deviations from

The M-SDUD provides amounts in prescriptions and units (e.g. grams, milliliters, pills, etc.). Earlier data shows some discrepancies in the number of prescriptions, likely contributing to reporting errors in South Dakota, Tennessee, and Washington (Table 1). Inspection of the year-to-year changes in prescriptions (e.g. Washington in 2006 was 141.5 fold higher than the prior year and 208.2 fold elevated relative to the subsequent year) reveals pronounced temporal instability. Further, the changes in prescriptions did not correspond with the number of units. After removing these extreme values, more than 142 million prescriptions and 9.7 billion units of oxycodone were distributed throughout the US from 2000-23. The average number of units per prescription was 66.0. The number of prescriptions and units of oxycodone peaked in 2012, with an increase of +218% and +183% respectively. The number of prescriptions decreased by −8% and the number of units by −17% from 2000-23. (Figure 1, Figure 3)

**Figure 3.**
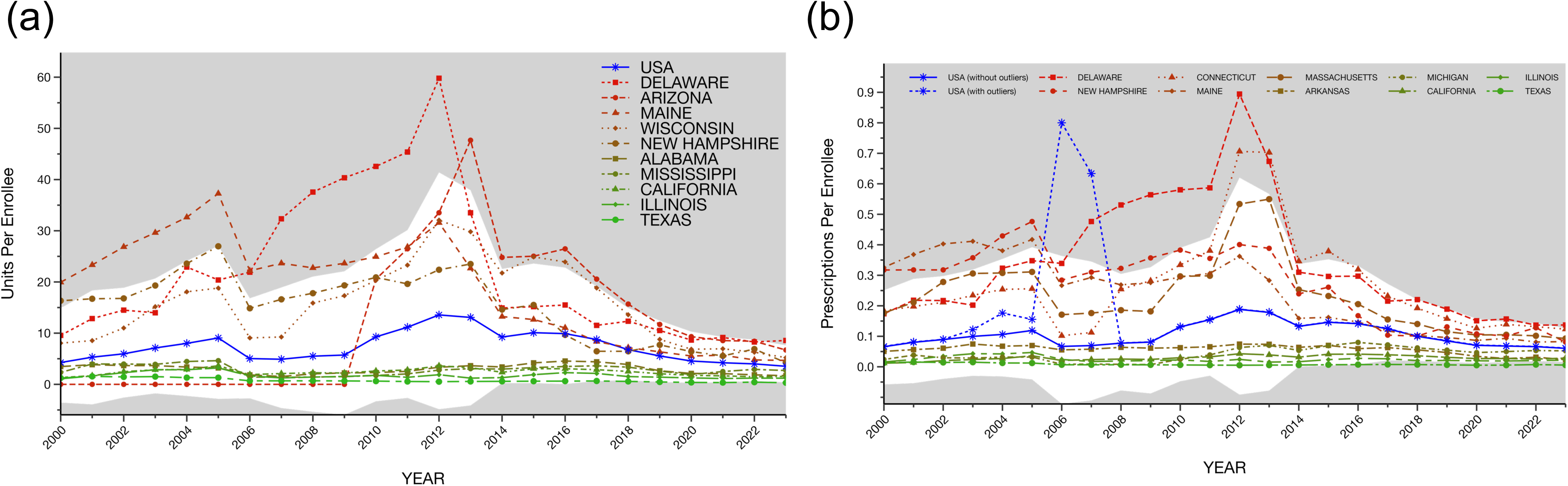
Oxycodone distribution for the top 5 and bottom 5 states as reported by the Medicaid State Drug Utilization Data in **(a)** units per enrollee and **(b)** prescriptions per enrollee. The shaded regions indicate ±1.96 standard deviations from the mean. “USA (without outliers)” for Prescriptions per Enrollee does not include the outlier values from South Dakota, Tennessee, and Washington from 2003-2007.

**Table 1.** Medicaid State Drug Utilization Data prescriptions, units, and total amount reimbursed for oxycodone from 2000-2010 for states with outliers likely due to reporting error. SD = South Dakota, TN = Tennessee, and WA = Washington. Concerning values for prescriptions are highlighted.

|  | 2000 | 2001 | 2002 | 2003 | 2004 | 2005 | 2006 | 2007 | 2008 | 2009 | 2010 |
| --- | --- | --- | --- | --- | --- | --- | --- | --- | --- | --- | --- |
| **State** | <u>Number of Prescriptions</u> |  |  |  |  |  |  |  |  |  |  |
| **SD** | 3,380 | 5,127 | 6,673 | 9,996 | 11,643 | 11,877 | 7,190 | 24,092,023 | 8,122 | 8,169 | 8,732 |
| **TN** | 27,131 | 78,053 | 110,227 | 253,252 | 2,914,201 | 1,535,862 | 2,871,497 | 144,252 | 166,723 | 189,800 | 205,960 |
| **WA** | 97,497 | 133,616 | 165,291 | 875,085 | 183,119 | 198,804 | 28,080,166 | 134,845 | 150,971 | 146,891 | 166,467 |
|  | <u>Number of Units</u> |  |  |  |  |  |  |  |  |  |  |
| **SD** | 174,109.00 | 270,852.00 | 366,071.00 | 588,262.00 | 672,352.00 | 713,270.00 | 465,628.00 | 249,553.00 | 539,571.00 | 535,938.00 | 576,888.00 |
| **TN** | 1,566,506.00 | 4,841,937.00 | 7,341,791.00 | 15,337,106.00 | 24,211,205.41 | 23,318,946.23 | 8,851,886.27 | 8,929,437.50 | 10,318,129.00 | 11,179,524.20 | 12,426,276.90 |
| **WA** | 6,342,380.00 | 8,737,138.47 | 10,665,915.44 | 12,647,823.56 | 12,992,987.00 | 14,709,182.50 | 8,983,049.00 | 9,878,594.07 | 11,346,947.50 | 10,485,891.00 | 12,238,654.80 |
|  | <u>Total Amount Reimbursed</u> |  |  |  |  |  |  |  |  |  |  |
| **SD** | 219,196.63 | 373,871.86 | 577,058.43 | 1,053,401.37 | 1,281,405.12 | 1,195,741.96 | 556,759.99 | 189,168,999.90 | 752,673.22 | 926,947.58 | 858,609.49 |
| **TN** | 2,044,642.01 | 8,143,864.66 | 13,851,081.67 | 28,355,311.77 | 70,892,324.64 | 105,243,425.11 | 285,308,844.46 | 9,019,127.10 | 10,378,039.61 | 11,748,616.63 | 11,694,600.21 |
| **WA** | 8,405,275.63 | 13,557,922.78 | 15,115,700.99 | 97,392,968.04 | 12,814,617.21 | 11,931,547.11 | 556,164,185.10 | 6,203,743.15 | 8,332,585.25 | 8,009,244.37 | 8,748,213.00 |

The M-PDP dataset provides the number of claims and 30-day fills for oxycodone from 2013-22. There were 182.2 million claims in total over these years. The number of beneficiaries with claims for the drug is also reported, allowing for a more accurate measure of per capita utilization. The pattern based on total enrollees (available for 2013-21) versus beneficiaries, which are available from the same dataset, differ greatly. Claims and 30-day fills per enrollee, which are more comparable to the above measures in ARCOS and Medicaid data, both peaked in 2015 with a +5% increase since 2013. Claims per enrollee decreased by −25%, and 30-day fills per enrollee decreased by - 26% between 2013 and 2021. Claims and 30-day fills per beneficiary peaked in 2020, increasing by +18% and +17%, respectively. They decreased by 2022, ending at +5% and +4% of their values from 2013. (Figure 1, Figure 4)

**Figure 4.**
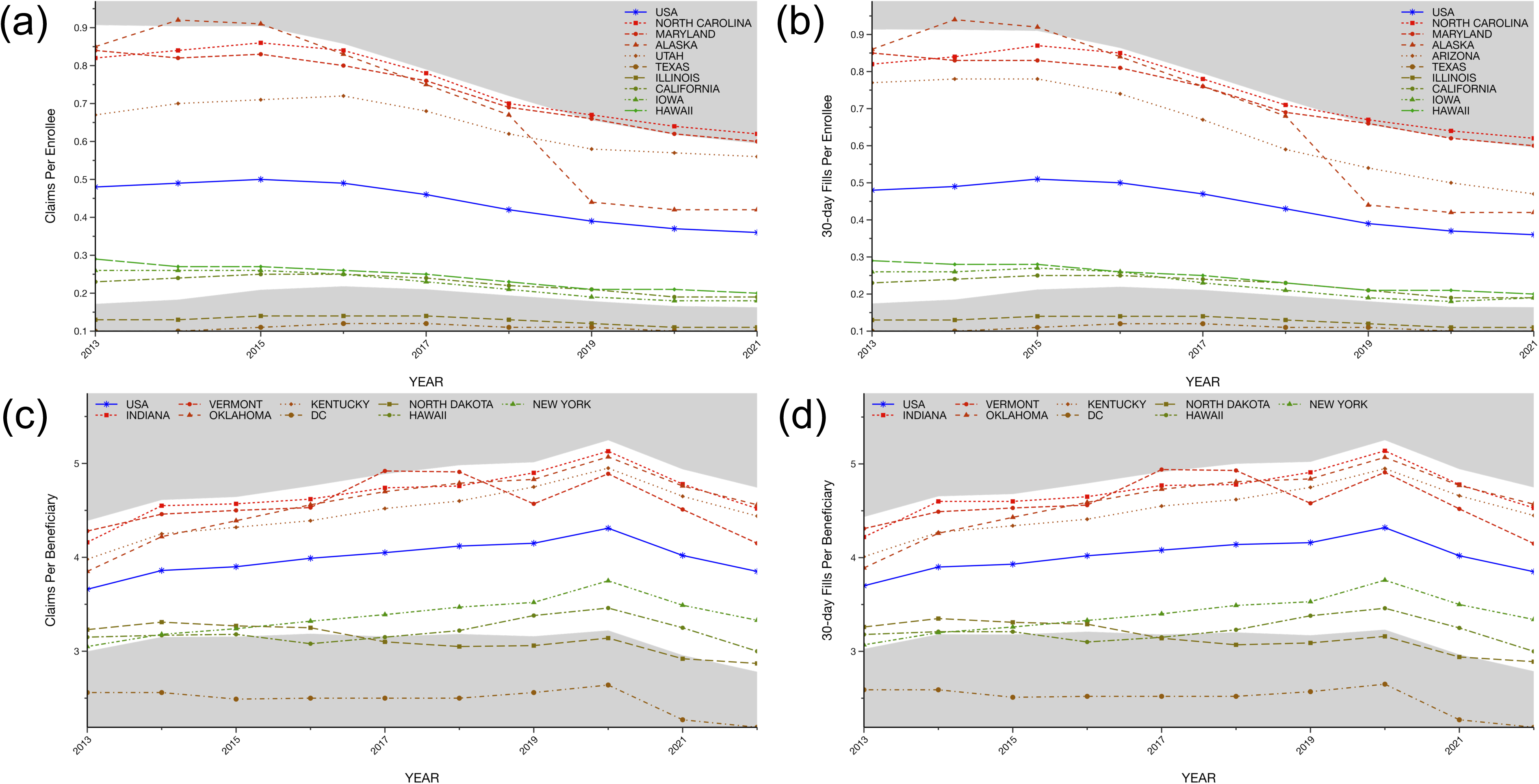
Oxycodone distribution for the top 5 and bottom 5 states as reported by the Medicare Part D Prescribers dataset in (a) units per total enrollees, (b) 30-day fills per total enrollees, (c) claims per beneficiaries with claims for oxycodone, (d) and 30-day fills per beneficiaries with claims for oxycodone. The shaded regions indicate ±1.96 standard deviations from the mean.

### Oxycodone Distribution by State

MME per person calculated using ARCOS data varied greatly between states in total and for each business type. The top and bottom states had an 8-fold difference in 2000 (Alaska = 177.7, Illinois = 23.4), a 15-fold difference at the peak in 2010 (Florida = 993.7, Texas = 66.7), and a 5-fold difference in 2023 (Tennessee = 223.3, Illinois = 42.6). Florida (2007-11), Delaware (2003-20), Tennessee (2012-21), Alaska (2000-02), and Missouri (2002-05) were distributing amounts of oxycodone greater than 1.96 standard deviations above the mean for persistent periods of time. Illinois (2012-23) and Texas (2012-23) were the only two states that were less than 1.96 standard deviations below the mean. (Figure 1, Figure 2)

Pharmacies, being the largest contributor to the overall trend, showed largely similar patterns between states. Florida (2007-11), Delaware (2003-20), Tennessee (2012-14, 2017-20), Alaska (2000-03), and Missouri (2001-05) were distributing substantially larger amounts of oxycodone per person than other states. Illinois (2013-18) and Texas (2013-18) were considerably lower than most states. State-level distribution patterns for hospitals differed greatly from the total and pharmacy groups. The District of Columbia (DC) (2000-11), Colorado (2008-23), Alaska (2000-01, 2008, 2010-23), and Montana (2000-03) had consistently high distribution per person compared to other states. Only Illinois (2000-02) ever fell below 1.96 standard deviations from the mean. The pattern for practitioners varied widely between states, but Florida (2000-10) stood in stark contrast from the others, peaking at more than 26,000 times greater MME per person in 2010 than the lowest state (Florida = 85.27, West Virginia = 0.0033). Similarly to the other business types, Alaska (2000-04) was elevated in the earlier years. Hawaii (2014-15, 2017-23) and Maryland (2014, 2018-19, 2021-23) have been distributing more oxycodone than other states more recently. (Figure 1, Figure 2)

Claims and prescriptions per enrollee in the M-SDUD also differed substantially between states. The top and bottom states by claims per enrollee had a 22-fold difference in 2000 (Alaska = 23.9, Illinois = 1.1), a 114-fold difference at the peak in 2012 (Delaware = 59.8, Texas = 0.5), and a 30-fold difference in 2023 (Maryland = 9.7, Texas = 0.3). Delaware (units: 2006-12; prescriptions: 2008-13) and Alaska (both units and prescriptions: 2000-02) were the only states to show a similar elevation as the ARCOS trends in both units and prescriptions per enrollee. Arizona (units: 2013-18; prescriptions: 2016), Maine (units: 2000-09; prescriptions: 2000-02), and Maryland (units: 2018-23; prescriptions: 2014-15, 2017-23) showed consistent elevations above 1.96 standard deviations from the mean. Texas (units: 2018, 2021-23; prescriptions: 2016-23) was the only state that fell below 1.96 standard deviations from the mean. The three states with abnormal prescription values (i.e., greater than 5*IQR+Q3) in the early years–South Dakota (2007), Tennessee (2004-06), and Washington (2003, 2006)– otherwise were similar to most states in their trends. Arizona was missing all data for oxycodone from 2000-09 because it did not participate in the Medicaid Drug Rebate Program until the Affordable Care Act was passed in 2010.^37^ (Figure 1, Figure 3)

The Medicare data spans a smaller timeframe, 2013-22, but has an interesting advantage in providing the number of beneficiaries that have claims for oxycodone. When looking at claims and 30-day fills per total Medicare Part D enrollees Illinois and Texas have substantially lower amounts for 2013-21 (2022 enrollment data is not yet available when data analysis was completed) while Alaska (2014-25), North Carolina (2020-21), and Tennessee (2015-21) have elevated amounts. However, when corrected by the number of beneficiaries, this effect disappears. The number of claims per beneficiary in DC is lower than 1.96 standard deviations from the mean for 2013-22, along with North Dakota (2017-21) and Hawaii (2016). When adjusting for number of beneficiaries, only Maine (2013) and Vermont (2017) ever go above 1.96 standard deviations from the mean for either claims or 30-day fills. For all states, the number of 30-day fills never differed from the number of claims by more than +2.18% (California in 2013). (Figure 1, Figure 4)

### Monetary Cost of Oxycodone

The Medicaid and Medicare data also offer the advantage of examining the cost associated with oxycodone. There was more than 8.7 billion dollars in reimbursement for oxycodone from 2000-23, more than 6.5 billion (74.7%) of which was paid by Medicaid. The total drug cost for Medicare was 14.75 billion dollars from 2013-22. Having the number of beneficiaries with claims for oxycodone allows a calculation of individual cost, which showed a peak of 384.56 dollars in 2014, with a gradual decline to 263.25 dollars per beneficiary in 2022.

### Correlations

ARCOS MME per person and M-SDUD claims per enrollee had a moderate positive correlation (*r* = 0.647, p < 0.001). ARCOS MME per person had a weak positive correlation with M-PDP claims per enrollee (*r* = 0.13, p = 0.005) and M-PDP claims per beneficiary (*r* = 0.271, p < 0.001). M-SDUD claims per enrollee was not associated with M-PDP claims per enrollee (*r* = 0.051, p = 0.275) and M-PDP claims per beneficiary (*r* = 0.029, p = 0.509). M-PDP claims per enrollee and claims per beneficiary had a non-significant positive correlation (*r* = 0.065, p = 0.116). (Figure 5)

**Figure 5.**
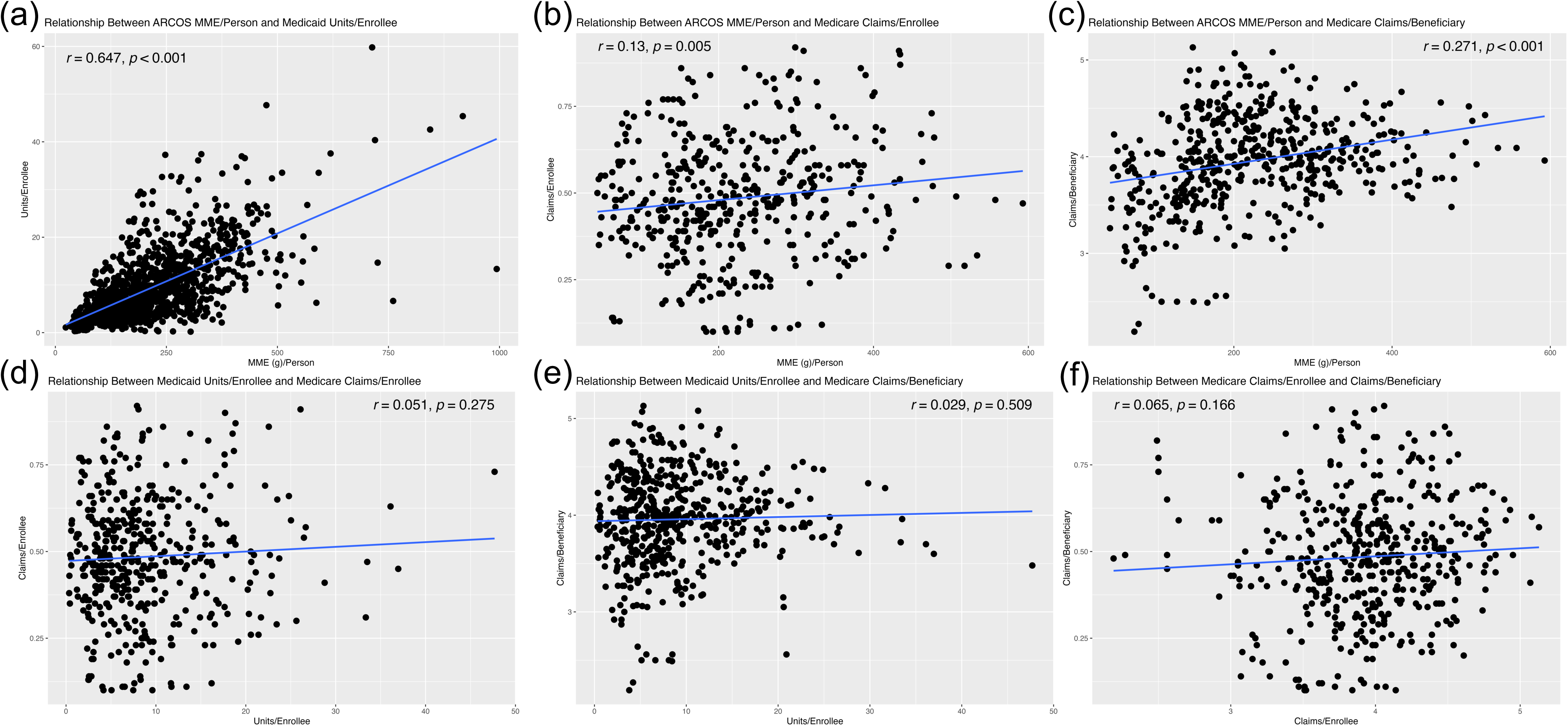
Scatter plots showing the relationships between **(a)** ARCOS MME/Person and Medicaid Units/Enrollee, **(b)** ARCOS MME/Person and Medicare Claims/Enrollee, **(c)** ARCOS MME/Person and Medicare Claims/Beneficiary, **(d)** Medicaid Units/Enrollee and Medicare Claims/Enrollee, **(e)** Medicaid Units/Enrollee and Medicare Claims/Beneficiary, and **(f)** Medicare Claims/Enrollee and Claims/Beneficiary. The Pearson correlation coefficient (*r*) and p-values are displayed on the respective plot.

## DISCUSSION

This study employed three complementary data sources–ARCOS, M-SDUD, and Medicare-PDP–to characterize oxycodone distribution throughout the US. ARCOS offers a comprehensive overview of national distribution, while Medicaid and Medicare datasets provide insights into prescribing trends among vulnerable populations. ARCOS provides weights of oxycodone distributed, Medicaid provides units and prescriptions, and Medicare provides claims and 30-day fills. Each source has advantages and disadvantages in ease of access/use and the type of information encompassed. ARCOS reports on distribution to 100% of the US population while in 2023 Medicaid covered 26% and Medicare covered 16%. Understanding these subtle differences can help healthcare professionals and policymakers refine opioid stewardship programs and enhance regulatory frameworks to curb misuse.

The ARCOS database revealed a notable increase in total oxycodone distribution from 2000-10, largely driven by pharmacies. Since the 2010 peak, oxycodone distribution has been on the decline, most likely due to regulatory actions including prescription drug monitoring programs and opioid prescribing limits. Differences between states were striking; Florida and Delaware had sharp increases with equally swift declines after the 2010 peak.^11,12,38^ Texas and Illinois had consistently lower distributions than other states.^39^ From 2000-23, the difference between the top and bottom states never dropped below five-fold extending a prior report.^40^ State-level disparities underscore regional differences in prescribing practices, healthcare infrastructure, and the effectiveness of policy interventions.

The M-SDUD showed a similarly large increase in oxycodone distribution but a later peak in 2012. Delaware stood out again with persistently elevated units and prescriptions in all years. New Hampshire, Massachusetts, and Maine also showed high numbers across many years but were not unusually elevated compared to other states in ARCOS. Texas and Illinois were again distributing considerably lower amounts of oxycodone. Notably, disproportionate and massive increases in prescription numbers in South Dakota, Tennessee, and Washington in the early 2000s are implausible, were removed, and call into question the integrity of the data in those earlier years. Although prior reports with M-SDUH have also identified occasional anomalous data, this is concerning for the veracity of reports that make use of this freely available dataset for a large portion of the US population.^17,27,28^

The M-PDP data showed a much different trend, possibly due to the population (individuals 65+ or with certain disabilities) or the timeframe covered by the data. From 2013-23, there was a decrease across most states in ARCOS and the M-SDUD with substantially less disparity between states. The Medicare database highlights the importance of the population being used to correct the data; trends across time for some states were considerably different depending on whether the claims/30-day fills were corrected by total enrollees or beneficiaries with claims for oxycodone. Both Illinois and Texas showed low claims and 30-day fills per enrollee, while Alaska, North Carolina, and Tennessee showed high numbers, but this was no longer the case when correcting for number of beneficiaries instead.

Correlations between the measures in each data source show that only ARCOS MME per person and M-SDUD units per enrollee had a moderate positive linear correlation (*r* = 0.647, p < 0.001). There may be more common factors driving the trends seen in these two measures compared with those influencing the changes in the M-PDP. The Medicare population is likely to have a higher comorbidity burden that can impact prescribing practices. The non-significant correlation within the M-PDP data between claims per enrollee and claims per beneficiary (*r* = 0.065, p = 0.166) highlights disparities when using a general vs. specific population.

The peaks and sharp declines in oxycodone distribution around 2010 are possibly due to a combination of regulatory programs, governmental interventions, and generally enhanced awareness of the risk of opioid dependence in healthcare by both patients and providers. Prescription monitoring programs and laws limiting the quantity dispensed for acute pain may have contributed to the decline we have seen. There is, however, mixed information about the effectiveness of laws in decreasing opioid prescriptions, and further work may better delineate factors that account for changes in oxycodone distribution.^41,42^ In 2010, there was a reformulation of OxyContin, a prominent brand name formulation, which made crushing the pill for nasal insufflation and dissolving the pill for intravenous injection more difficult.^43^ Some research shows that this change may in turn decrease the misuse potential of, and demand for, oxycodone.^44,45^ However, it is important to appreciate that according to the Treatment Episode Data Set, the predominant route of administration for those that were admitted to treatment programs for prescription opioids was oral.^46^ Around the same time, there was increasing legal action against physicians who inappropriately prescribe controlled medications. These legal efforts were prominent in Florida and extended to other states in following years.^47^

All three databases share some strengths and weaknesses. They are all publicly accessible (unlike IQVIA) and deidentified for examination. However, all reported values pertain to licit distribution and any diversion of drugs to a third party cannot be determined. Similarly, information about whether, or how much of, the drug was used by recipients is unavailable.

ARCOS is a comprehensive data source that has data over a large time frame and is the only source that readily quantifies the amount of oxycodone distributed. It has advantages over the less complete IQVIA database, as ARCOS covers Veteran’s Affairs, Indian Health Services facilities, hospitals, and independent pharmacies. The data includes all transactions and shipments between distributors and may result in overestimation. Some amount of the oxycodone reported in pharmacy, practitioners, and teaching institutions business activities may be used by veterinarians.^48^ The American Community Survey data used to normalize for population is limited by counting of undocumented individuals and may be an underestimate.^49^

The data is also difficult to transform into an analyzable format. The ARCOS data released by the Washington Post is easy to work with but initially only covered 2006-14.^50^ The reports are provided as PDF files with inconsistent formats across years. Transferring the tables into a filetype that is conducive to analytic work (i.e. .csv) is most easily accomplished with an online conversion website, which can lead to errors. Utilizing the packages tabula-py and pandas (RRID:SCR_018214) for Python allowed for a more reproducible approach with manual accuracy checks.

The M-SDUD is easily accessible online with the ability to filter and download data. Providing the number of prescriptions is useful to put the distribution in a clinical context. It additionally provides the associated financial costs, which is important for pharmacoeconomists. The study population is large but limited compared to ARCOS. The Medicaid data has a lower spatial resolution, providing state-level data compared to ARCOS data which can be grouped by zip code. It is also not possible to easily quantify the amount of oxycodone, since there are a multitude of different formulations reported and the NDC units code is less easily conducive to a systematic conversion to weight or volume.

The broad range of the provided drug names within and across the generic and brand names (i.e. oxycodone, oxycodone hcl, oxycodone hydrochloride; Percocet, Percocet 5, Percocet|oxycodone) makes it difficult to effectively filter the data to capture all formulations. Additionally, it is difficult to get a list of all NDC codes for oxycodone products over the entire range of data as they can change between years. Although some small amounts may still be missed, using the NDC code directory and searching the data for known patterns using regular expressions (character sequences that define a match pattern) provides a more comprehensive extraction of data.

The M-PDP data is also easily accessible online and the most user friendly. It is simple to filter the data based on either generic or brand names. There are also flags for opioids and long-acting opioids (as well as antipsychotics and antibiotics) for the study of these classes of drugs. Data on how many beneficiaries had claims for each drug allows the calculation of the most accurate measure of oxycodone distribution per person. Uniquely, the M-PDP data can also provide information about individual providers, though data is broadly grouped into categories like opioids. Like Medicaid, this dataset also reveals the financial costs but has a spatial resolution limited to states and a population that only includes those enrolled in Medicare Part D, an optional benefit that covers prescription drugs. The specific amounts of oxycodone are also not quantifiable as there is no information about each formulation. This dataset is the smallest and spans only 2013-22, which misses the peak years based on the other data sources.

By analyzing multi-source data, clinicians and policy makers can develop more effective interventions to mitigate opioid misuse while ensuring appropriate access to pain relief. As data collection and sharing continues to improve, ARCOS, M-SDUD, and M-PDP data can be valuable resources for monitoring drug utilization across the US. Interpreting trends in prescription opioid use at the state-level can provide insight into the effectiveness of guidelines and regulations or reveal utilization trends that require intervention. The results of this report can aid the efforts to limit prescription opioid misuse, and it also lays the foundation for future works to leverage these data sources to look closely at states of interest. The findings reported here may also be valuable internationally for other countries looking to avoid the opioid excesses of the US.

## Data Availability

All data produced in the present study are available upon reasonable request to the authors.

https://github.com/solgamaj/ARCOS-MEDICAID-MEDICARE

## Acknowledgements

Jove Graham, PhD and Mellar Davis, MD are appreciated for their mentorship during the project. The Geisinger Commonwealth School of Medicine Summer Research Immersion Program is appreciated for providing a stipend and supporting the project during the summer of 2022.

## Data Sources

ARCOS data is publicly accessible at https://www.deadiversion.usdoj.gov/arcos/retail_drug_summary/arcos-drug-summary-reports.html.

Medicaid State Drug Utilization Data is Publicly Accessible at https://www.medicaid.gov/medicaid/prescription-drugs/state-drug-utilization-data/index.html.

Medicare Part D Prescribers data is publicly accessible at https://data.cms.gov/provider-summary-by-type-of-service/medicare-part-d-prescribers.

Scripts and data used for this study can be found at https://github.com/solgamaj/ARCOS-MEDICAID-MEDICARE.

